# Genomic recombination of rapidly evolving mpox Ib strains compounds the challenges of the 2024 outbreak

**DOI:** 10.1101/2024.09.18.24313912

**Authors:** Ting-Yu Yeh, Patrick J. Feehley, Michael C. Feeley, Chieh-Fan Chen, Tung-Yuan Tsai, Hsiang-Lan Cheng, Gregory P. Contreras

**Author notes:** Corresponding author: Ting-Yu Yeh, MD, Auxergen Inc., Rita Rossi Colwell Center, 701 E Pratt Street, Room 4010, Baltimore, MD 21202, USA. These authors contributed equally (Patrick J. Feehley, Michael C. Feeley, Gregory P. Contreras).

## Abstract

The World Health Organization recently declared the 2024 mpox virus (MPXV Ib) outbreak a public health emergency of international concern. We report that in 2023-2024, MPXV clade Ib genomes are diverging at a faster rate than clade IIb (2022), primarily due to an unusually high incidence of recombination. Phylogenetic analysis revealed that Ib strains have diverged into four lineages, and they have evolved into 14 subgroups based on nine tandem repeat (TR) polymorphisms. These findings confirms that TRs in MPXV Ib are mutating at a significantly higher frequency compared to the 2022 outbreak (clade IIb, 11 subgroups). Linkage disequilibrium analysis also identified 10 recombination clusters among all 4 lineages, with recombination incidence in Ib being twice as high as in IIb. This suggests that a higher rate of superinfection is contributing to ongoing recombination among populations infected with clade Ib. Prompt action is necessary to prevent the emergence of more lethal mpox strains.

## Introduction

On August 14, 2024, the World Health Organization declared the mpox virus (MPXV Ib) outbreak a public health emergency of international concern with a three percent fatality rate—18,737 mpox cases and 541 deaths across 14 countries — reported as of August 16, 2024. ^1^ The European CDC has suggested the outbreak size may be larger than reported due to under-ascertainment and under-reporting. ^2^ Here we report that MPXV clade Ib genomes in 2024 are diverging faster than clade IIb (2022) due to an unusually higher incidence of recombination. Our data underscores the urgency required to reduce human morbidity.

## Methods

Genomic sequences of MPXV Ib from October 13, 2023 to August 28, 2024 (excluding low coverage, N=32) were obtained from GASAID (www.gasaid.com). Phylogenetic, tandem repeat (TR), and the linkage disequilibrium (LD) analyses were performed as previously described.^4^ Subgroups were categorized by (1) the presence of TR B; and (2) the TR numbers of TR A, D, E, G, and H. HaploView was used to display LD, haplotype blocks, and recombination by plotting 95% confidence bounds for *D*’ (the normalized values of the coefficient of LD). ^4^

## Results

It has been suggested that the APOBEC3 mutation in new clade I strain contains could lead to faster viral evolution.^3^ However, how this influences the MPXV Ib genome remains unclear. We examined 32 MPXV Ib sequences worldwide in GISAID. Alignment-based phylogenetic analysis showed that Ib strains have diverged into 4 lineages (Figure 1A). Analyzing tandem repeats (TRs, Figure 1B), an alignment-free method more resistant to sequencing error,^4^ revealed that Ib sequences have evolved into 14 subgroups based on 9 TR polymorphisms (219-738 base pairs) (Figure 1C). This result confirms that the MPXV Ib TRs are mutating much more frequently compared to the 2022 pandemic (clade IIb, 11 subgroups) .^4^

**Figure 1.**
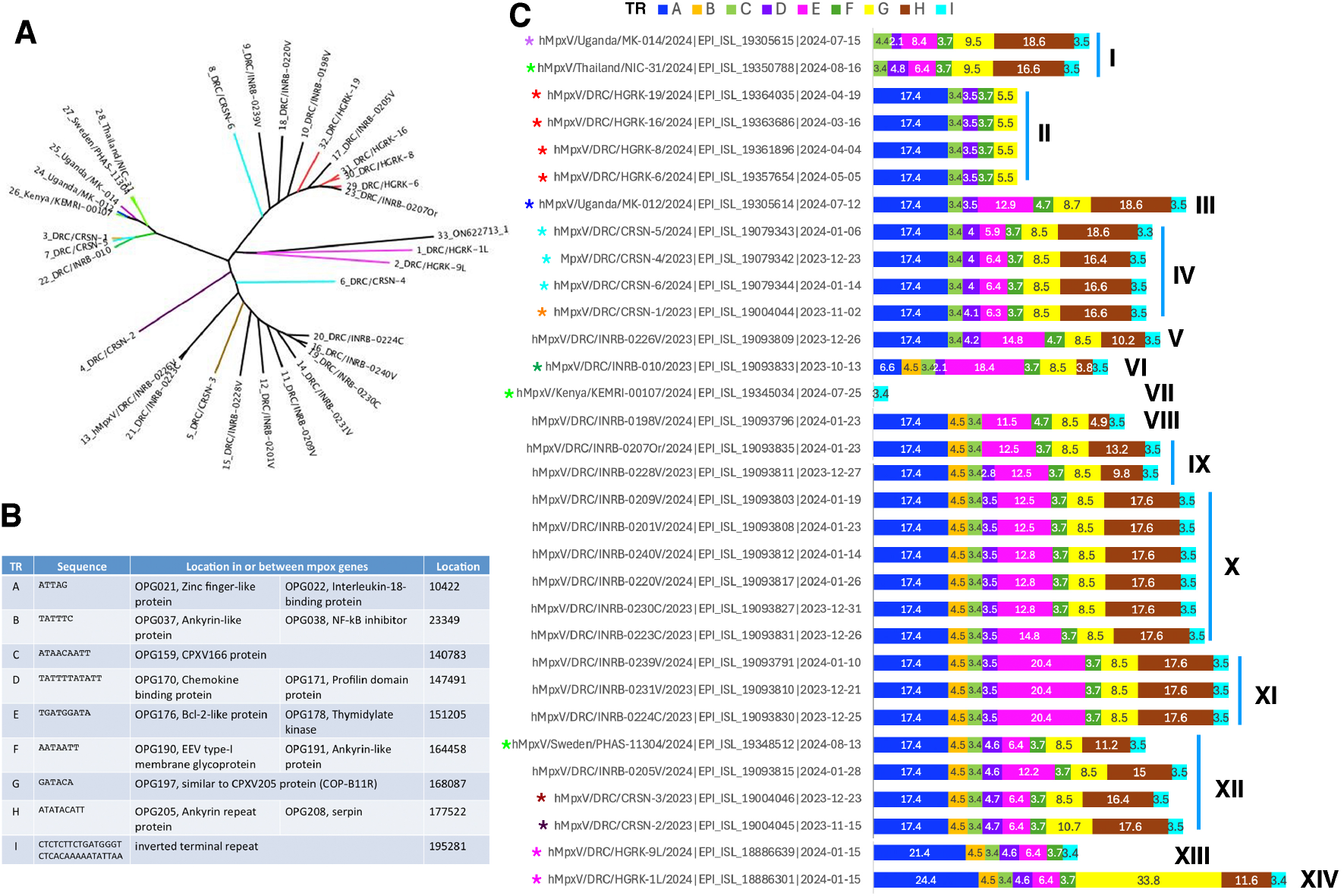
(A) Rooted phylogenetic tree of MPXV Ib genomes, October 13, 2023 to August 28, 2024 (N=32). Sequence alignment was generated using MAFFT 7 and the phylogenetic tree was visualized using FigTree. Rooting was done by introducing Belgium MPXV REGA2 (accession number ON622713) as an outgroup virus. Recombinants are labeled in color. (B and C) Tandem repeats (TR, A to I) among mpox clade Ib sequences in 2024 endemics. (B) Characterization and location of TRs. (C) TR numbers are present in the boxes to detect TR polymorphisms and to categorize 14 different subgroups. Ten recombinant clusters from Figure 2 are labeled with color asterisks.

We and others reported the first MPXV recombination by natural transmission in the 2022 pandemic.^4,5^ In 2024, however, MPXV Ib genomes appear to recombine much more quickly. LD analysis detected 10 independent recombination clusters (Figure 2B-E). Recombinant sequences include DRC IRB010, Thailand NIC31/Sweden PHAS11304/Kenya KEMRI00107, DRC CRSN-4/5/6, DRC CRSN-1, 2, 3; DRC HGRK1L/9L; DRC HGRK6/8/16/19, Uganda MK012; and Uganda MK014 (Figure 2, asterisks). High recombination rate was also detected by haplotype block analysis between nucleotide 144961/148651 and 166442/166579 (multi-allelic *D*’ statistics: 0.94 and 0.98, respectively) (Figure 2D). TR analysis also showed that DRC INRB0205V exhibited recombination properties (Figure 1C). These results indicate that MPXV Ib recombination has occurred as early as October 2023 and is present in all 4 lineages now (Figure 1A). Compared to IIb (705 recombinants of 2526 sequences) ^5^, Ib recombination incidence is two-fold higher (56.3% versus 27.9%). This suggests that a higher rate of superinfection is leading to recombination and is ongoing among Ib-infected populations.

**Figure 2.**
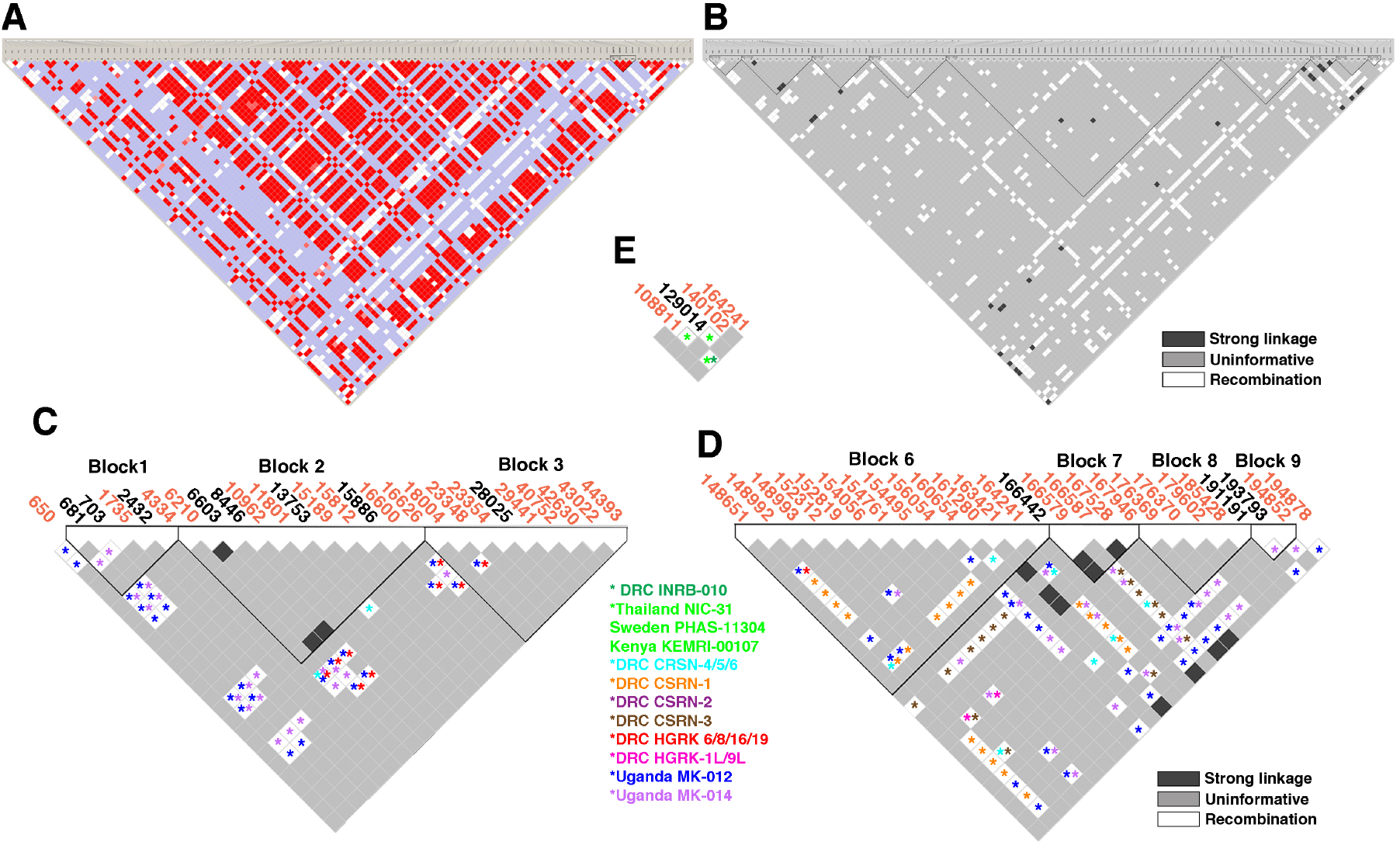
Linkage disequilibrium (LD) analysis of MPXV clade Ib polymorphisms. (A) HaploView (Broad Institute; Cambridge, MA) LD heat map among 115 sites of Ib polymorphism. Red squares indicate high levels of LD, and white and blue squares represent low levels of LD. Color scheme is described in detail previously.^4^ (B to E) Haplotype blocks organization to display the confidence bounds color scheme. The location of single nucleotide (black) or deletion (orange) polymorphism are based on the Sweden PHAS11304 sequence and listed at the top. The haplotype blocks of (B) (block 1 to 3) and (C) (block 6 to 9) represent the solid spine of strong LD. Strong LD (31 associations, black squares) is defined if the upper 95% confidence bound for *D*’ is above 0.98 and the lower bound is above 0.7. Recombination (636 associations, white squares) is defined if pairs for which the upper confidence bound of *D*’ is below 0.9. Recombination plots of the full length (B), the 5’ end (C, nucleotide 1-44393), 3’ end (D, nucleotide 148651-194878), and (E) nucleotide 108811-164241 of MPXV Ib genome are shown. Recombination clusters are labeled with color asterisks.

## Discussion

Even with limited sequence availability, our findings confirmed that MPXV Ib has generated more new recombinant variants and subgroups than IIb over the entire 2022 outbreak. The lack of laboratory capacity hinders the confirmation of mpox cases in many African countries.^1^ This results in a lack of available MPXV Ib sequences. It is therefore reasonable to conclude that the mutation and recombination rate of MPXV Ib is very likely underestimated. It has also been shown previously that poxvirus recombinants can arise in virus-by-virus crosses within a genus (e.g., variola-by-rabbitpox).^6^ Therefore, action should be taken immediately to avoid outbreaks of potentially more lethal strain. Given the fact that new variants are still emerging, best practices of genomic surveillance and enhanced transparency of MPXV sequences would enable a more thorough analysis, providing critical insights to the medical and scientific community about the 2024 outbreak.

## Data Availability

All data produced in the present study are available upon reasonable request to the authors

https://www.gasaid.com

## Ethical approval

None declared.

## Author Contributions

Yeh and Contreras had full access to all of the data in the study and take responsibility for the integrity of the data and the accuracy of the data analysis.

Concept and design: Yeh

Acquisition, analysis, or interpretation of data: Yeh, PJ Feehly, MC Feehly, Chen, Tsai, Cheng

Drafting of the manuscript: Yeh, PJ Feehly, MC Feehly, Contreras

Critical revision of the manuscript for important intellectual content: Yeh, Chen Statistical analysis: Yeh

Supervision: Yeh, Contreras

## References

1. Adepoju P. Mpox declared a public health emergency. Lancet. 2024; 404(10454): E1–E2. doi: 10.1016/S0140-6736(24)01751-3.

2. European Centre for Disease Prevention and Control. Risk assessment for the EU/EEA of the mpox epidemic caused by monkeypox virus clade I in affected African countries – 16 August 2024. ECDC: Stockholm; 2024.

3. Vakaniaki EH, Kacita C, Kinganda-Lusamaki E, et al. Sustained human outbreak of a new MPXV clade I lineage in eastern Democratic Republic of the Congo. Nature Med. 2024; 10.1038/s41591-024-03130-3

4. Yeh TY, Hsieh ZY, Feehley MC, et al.; Recombination shapes the 2022 monkeypox (mpox) outbreak. Med. 2022; 3(12):824–826. doi: 10.1016/j.medj.2022.11.003

5. Alfonsi T, Bernasconi A, Chiara M, Ceri S. Data-driven recombination detection in viral genomes. Nat Commun. 2024; 15:3313. doi.org/10.1038/s41467-024-47464-5

6. Evans DH. Poxvirus recombination. Pathogens. 2022; 11(8):896. 10.3390/pathogens11080896

